# State-Level Excess Mortality in US Adults During the Delta and Omicron Waves of COVID-19

**DOI:** 10.1101/2023.03.07.23286933

**Authors:** Benjamin Renton, Chengan Du, Alexander Junxiang Chen, Shu-Xia Li, Zhenqiu Lin, Harlan M. Krumholz, Jeremy Samuel Faust

## Abstract

**Introduction:** The US has continued to see excess mortality through the Delta and Omicron periods. We sought to quantify excess mortality on a state level and calculate potential deaths averted if all states matched the excess mortality rates of those with the 10 lowest excess mortality rates.

**Methods:** Observational cohort, US and state-level data. Expected monthly deaths were modeled using pre-pandemic US and state-level data (2015-2020). Mortality data was accessed from CDC public reporting.

**Results:** We find that during the Delta and Omicron waves, the US recorded over 596,000 excess deaths. 60% of the nation’s total excess mortality during these periods could have been averted if all states had excess mortality rates equal to those with the 10 lowest excess mortality rates.

**Conclusion:** With large differences in excess mortality across US states in this 15-month study period, we note that a significant portion of deaths could have been averted with higher vaccination rates, improved mitigation policies, and adherence to other behaviors.

## Introduction

The US has recorded substantial all-cause excess mortality during the COVID-19 pandemic.^1^ States had varying policies and strategies to address the pandemic. States also varied in their excess mortality, a metric that takes into account baseline health risks by determining how their pandemic mortality compared with their predicted mortality. While vaccination rates likely played an important role, other characteristics likely also influenced any detected differences.^2,3^

Accordingly, we quantified state-level excess deaths and estimated the number of excess deaths that could have potentially been averted if all US states had excess death rates matching the 10 US states with the lowest excess death rates during the SARS-CoV-2 variant era (i.e., the Delta and Omicron periods).

## Methods

We applied seasonal autoregressive integrated moving average (sARIMA) models to monthly all-cause death data from the Centers for Disease Control and Prevention and state population counts (2014-2020) to create monthly population and expected death estimates for the study period (July 1, 2021-September 30, 2022), correcting for population losses due to cumulative pandemic-associated excess deaths.^4,5^ Excess deaths were defined as observed minus expected deaths.

For each state, we modeled three age groups of adults (18-49, 50-64, and ≥65 years), which were then combined, creating composite excess mortality estimate. The study assessed five three-month periods based on dominant variants in sequence surveillance; July 1-September 30, 2021 (Early Delta), October 1-December 31, 2021 (Late Delta/Early Omicron), January 1-March 31, 2022 (Omicron BA.1), April 1-June 30, 2022 (Omicron BA.2), July 1-September 30, 2022 (Omicron BA.5). For each period and age group, we determined the ten states with the lowest and highest excess mortality. The number of potential excess deaths averted (had all states recorded excess mortality rates observed among the ten with the lowest rates) was computed for each period and age group, and summed for an all-US adult estimate.^6,7^

Analyses were conducted with R version 4.0.3. This study used publicly available data and was not subject to institutional review approval per HHS regulation 45-CFR-46.101(c).

## Results

During the Delta and Omicron periods, the US observed 596,173 (95% CI 514,861-677,486) excess deaths. The ten US states with the highest excess mortality had a combined excess mortality rate over five times the rate of the ten lowest excess mortality states (492.1 versus 93.9 per 100,000, Table). Had all states matched the excess death rate of the ten lowest excess mortality states, an estimated 356,009 excess deaths nationwide could potentially have been averted during the period, representing 60% of the nation’s total contemporaneous excess mortality. Excess deaths per 100,000 were higher during Early Delta (67.9), Late Delta/Early Omicron (61.9), and Omicron BA.1 (62.5) than during Omicron BA.2 (13.3) and Omicron BA.5 (24.7) (Table).

Excess deaths per 100,000 people varied widely among states, from 80.6 per (North Dakota) to 479.8 (West Virginia) overall and within periods (Table). Among states with higher excess mortality than the 10 lowest excess mortality states, the percent of excess deaths that could have potentially been averted ranged from 36% (Maryland) to 78% (West Virginia).

## Discussion

There were vast differences in excess mortality across US states during from July 2021-September 2022. By using the top quintile of US states as best-case scenario benchmarks, estimates for deaths potentially averted represent results that would be achievable in other states. Vaccination rates may explain some of the observed differences (Table), but jurisdiction-level policies and other behaviors also likely contributed.

Strengths of this observational study include using excess mortality rather than COVID-19 mortality, which provides better assessments of state-level performance by minimizing coding biases and including all deaths potentially modified by the pandemic. In addition, the focus on excess mortality (i.e., changes from baseline expected deaths), inherently takes existing risk differences across states into account. Study limitations include use of provisional mortality data and lack of information on causality.

## Data Availability

All data produced in the present study are available upon reasonable request to the authors

## Author Contributions

Dr. Faust and Mr. Renton had full access to all of the data in the study and takes responsibility for the integrity of the data and the accuracy of the data analysis.

*Concept and design:* Faust, Du, Krumholz, Renton.

*Acquisition, analysis, or interpretation of data:* All authors.

*Drafting of the manuscript:* Renton, Faust.

*Critical revision of the manuscript for important intellectual content:* All authors.

*Statistical analysis:* Faust, Du, Li, Lin, Renton.

*Administrative, technical, or material support:* Faust, Du, Lin.

*Supervision:* Faust, Krumholz.

*Data visualization:* Renton.

## Conflict of Interest Disclosures

Dr. Krumholz reported receiving consulting fees from UnitedHealth, Element Science, Aetna, Reality Labs, F-Prime, and Tesseract/4Catalyst; serving as an expert witness for Martin/Baughman law firm, Arnold and Porter law firm, and Siegfried and Jensen law firm; being a cofounder of Hugo Health, a personal health information platform; being a cofounder of Refactor Health, an enterprise health care, artificial intelligence-augmented data management company; receiving contracts from the Centers for Medicare & Medicaid Services through Yale New Haven Hospital to develop and maintain performance measures that are publicly reported; and receiving grants from Johnson & Johnson outside the submitted work. No other disclosures were reported.

**Table.**
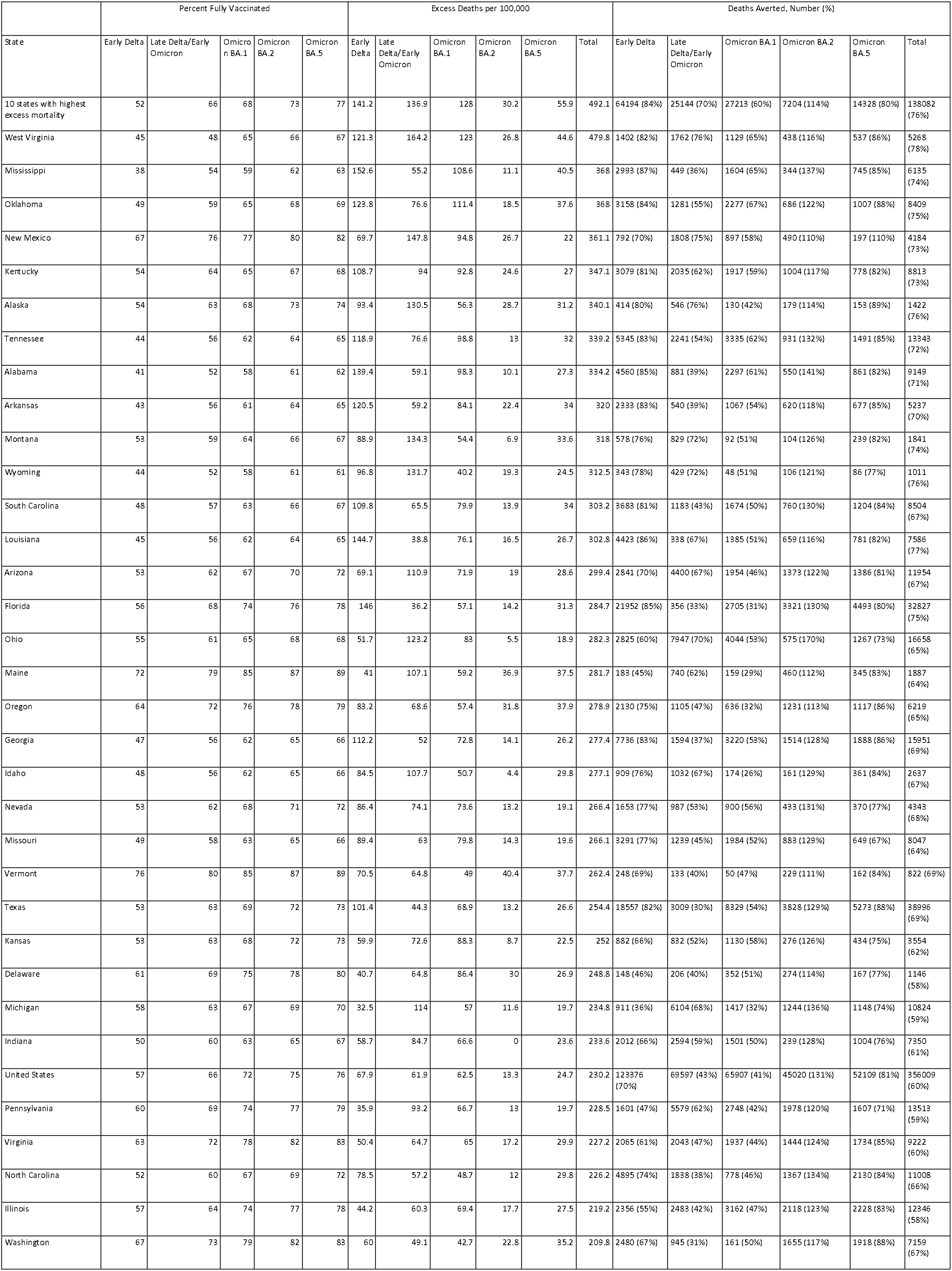
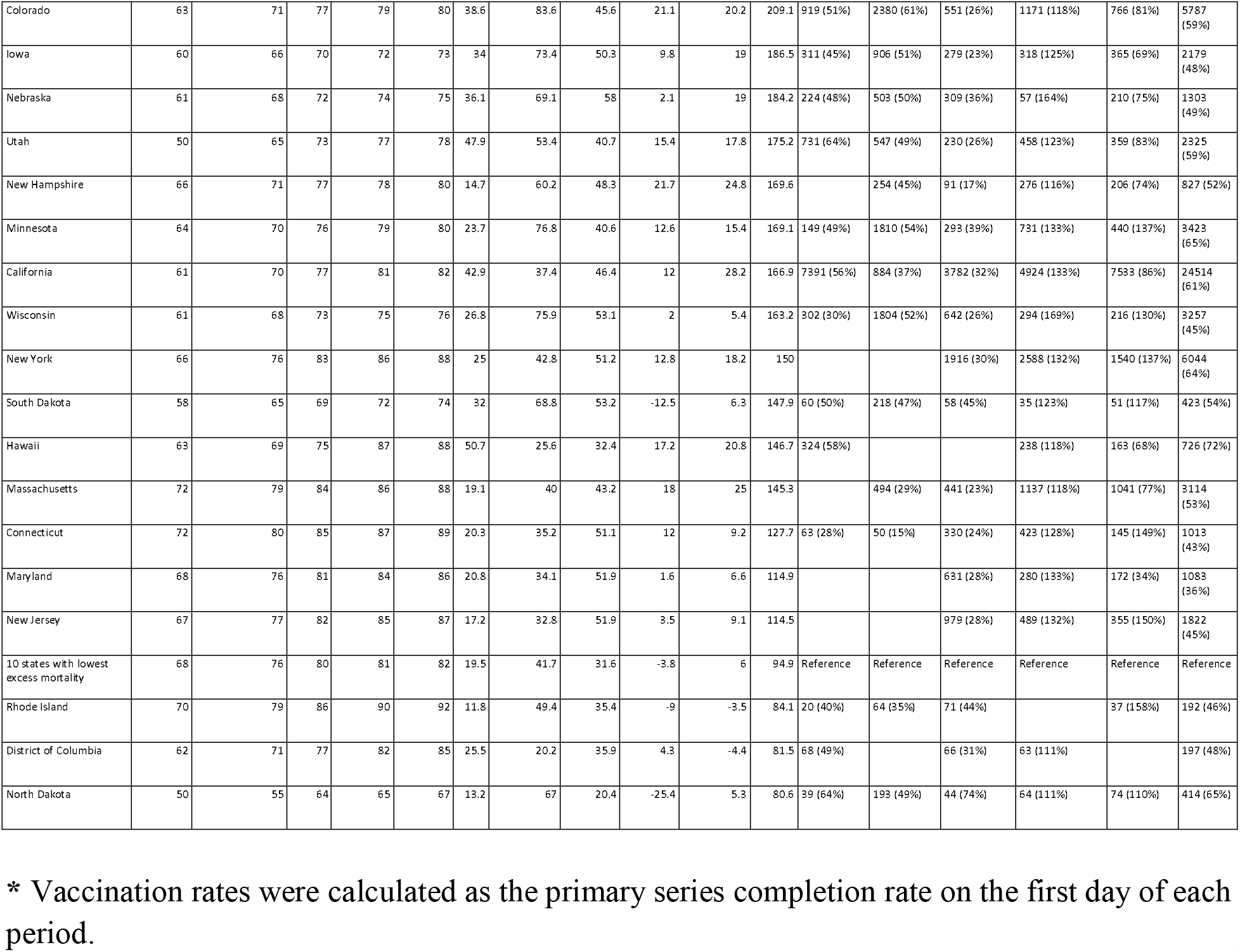

